# Accuracy of Computable Phenotyping Approaches for SARS-CoV-2 Infection and COVID-19 Hospitalizations from the Electronic Health Record

**DOI:** 10.1101/2021.03.16.21253770

**Authors:** Rohan Khera, Bobak J. Mortazavi, Veer Sangha, Frederick Warner, H. Patrick Young, Joseph S. Ross, Nilay D. Shah, Elitza S. Theel, William G. Jenkinson, Camille Knepper, Karen Wang, David Peaper, Richard A Martinello, Cynthia A. Brandt, Zhenqiu Lin, Albert I. Ko, Harlan M. Krumholz, Benjamin D. Pollock, Wade L. Schulz

## Abstract

**Objective:** Real-world data have been critical for rapid-knowledge generation throughout the COVID-19 pandemic. To ensure high-quality results are delivered to guide clinical decision making and the public health response, as well as characterize the response to interventions, it is essential to establish the accuracy of COVID-19 case definitions derived from administrative data to identify infections and hospitalizations.

**Methods:** Electronic Health Record (EHR) data were obtained from the clinical data warehouse of the Yale New Haven Health System (Yale, primary site) and 3 hospital systems of the Mayo Clinic (validation site). Detailed characteristics on demographics, diagnoses, and laboratory results were obtained for all patients with either a positive SARS-CoV-2 PCR or antigen test or ICD-10 diagnosis of COVID-19 (U07.1) between April 1, 2020 and March 1, 2021. Various computable phenotype definitions were evaluated for their accuracy to identify SARS-CoV-2 infection and COVID-19 hospitalizations.

**Results:** Of the 69,423 individuals with either a diagnosis code or a laboratory diagnosis of a SARS-CoV-2 infection at Yale, 61,023 had a principal or a secondary diagnosis code for COVID-19 and 50,355 had a positive SARS-CoV-2 test. Among those with a positive laboratory test, 38,506 (76.5%) and 3449 (6.8%) had a principal and secondary diagnosis code of COVID-19, respectively, while 8400 (16.7%) had no COVID-19 diagnosis. Moreover, of the 61,023 patients with a COVID-19 diagnosis code, 19,068 (31.2%) did not have a positive laboratory test for SARS-CoV-2 in the EHR. Of the 20 cases randomly sampled from this latter group for manual review, all had a COVID-19 diagnosis code related to asymptomatic testing with negative subsequent test results. The positive predictive value (precision) and sensitivity (recall) of a COVID-19 diagnosis in the medical record for a documented positive SARS-CoV-2 test were 68.8% and 83.3%, respectively. Among 5,109 patients who were hospitalized with a principal diagnosis of COVID-19, 4843 (94.8%) had a positive SARS-CoV-2 test within the 2 weeks preceding hospital admission or during hospitalization. In addition, 789 hospitalizations had a secondary diagnosis of COVID-19, of which 446 (56.5%) had a principal diagnosis consistent with severe clinical manifestation of COVID-19 (e.g., sepsis or respiratory failure). Compared with the cohort that had a principal diagnosis of COVID-19, those with a secondary diagnosis had a more than 2-fold higher in-hospital mortality rate (13.2% vs 28.0%, P<0.001). In the validation sample at Mayo Clinic, diagnosis codes more consistently identified SARS-CoV-2 infection (precision of 95%) but had lower recall (63.5%) with substantial variation across the 3 Mayo Clinic sites. Similar to Yale, diagnosis codes consistently identified COVID-19 hospitalizations at Mayo, with hospitalizations defined by secondary diagnosis code with 2-fold higher in-hospital mortality compared to those with a primary diagnosis of COVID-19.

**Conclusions:** COVID-19 diagnosis codes misclassified the SARS-CoV-2 infection status of many people, with implications for clinical research and epidemiological surveillance. Moreover, the codes had different performance across two academic health systems and identified groups with different risks of mortality. Real-world data from the EHR can be used to in conjunction with diagnosis codes to improve the identification of people infected with SARS-CoV-2.

## BACKGROUND

The COVID-19 pandemic has led to the rapid adoption of real-world evidence to guide the treatment of and the public health response to a novel pathogen.[1-5] The identification of both SARS-CoV-2 infection and COVID-19 hospitalization is of current clinical and regulatory importance given the need for case identification for epidemiologic surveillance to track the infections, mortality, and vaccine effectiveness. Similarly, clinical predictive models that rely on appropriate case classification and studies that track the long-term effects of SARS-CoV-2 infection may be biased if case definitions are inaccurate or capture only subsets of individuals infected with SARS-CoV-2. Administrative data represent a widely available real-world data source to monitor COVID-19 cases and hospitalizations using diagnosis codes.

Administrative data, a source of real-world data (RWD) generated from billing claims, can be used for disease surveillance, to follow hospitalization rates, and characterize patient outcomes on a large scale as well as evaluate the effects of health policy for these measures.[6-10] However, reliance on claims alone may lead to erroneous inferences, as has been shown for other conditions.[11 12] To ensure that high-quality data guide national policy and biomedical research, there is a need to evaluate the accuracy of the diagnostic code-based approaches used to define cases of SARS-CoV-2 infection and hospitalization.

The adoption of health information technology systems has positioned health systems to improve case identification by incorporating more detailed clinical data from the electronic health record (EHR) with diagnosis codes, which allows for the development of more accurate computable phenotypes.[13-18] The EHR represents a potential advance over the use of administrative data alone for case identification, outcome ascertainment, and validation of computable phenotyping approaches.

In this study from two large health systems with academic and community-based practices, we evaluate the accuracy of various approaches to identify people with SARS-CoV-2 infection and COVID-19 hospitalizations based on diagnostic codes and laboratory testing results from the EHR. We also assess the impact of cohort definition on the ascertainment of in-hospital mortality rates for COVID-19.

## METHODS

### Data Sources

We used EHR-derived data from Yale New Haven Health System (Yale), a large academic health system consisting of 5 distinct hospital delivery networks and associated outpatient clinics located in Connecticut and Rhode Island. Data from our EHR clinical data warehouse were transformed into the Observational Medical Outcomes Partnership (OMOP) common data model (CDM) using our computational health platform.[13 19] We used a versioned extract of the OMOP data from March 3, 2021 and analyzed testing and discharge information from April 1, 2020, when the COVID-19 specific *International Classification of Diseases-10*^*th*^ *Edition-Clinical Modification (*ICD-10-CM) diagnosis was introduced, through March 1, 2021. Admissions were limited to those with a visit start date before January 31, 2021 to allow for a majority of those admitted to have been discharged.[3]

To evaluate the generalizability of our observations, a similar cohort was constructed in the three hospital delivery networks of the Mayo Clinic. Mayo Clinic is an academic health system headquartered in Rochester MN, with two additional destination medical centers in Phoenix, AZ, Jacksonville, FL, and several regional and critical access hospitals in Minnesota and Wisconsin.

The study was approved by the Institutional Review Boards of Yale University and Mayo Clinic. Data were independently analyzed at each site.

### Cohort Definitions

#### SARS-CoV-2 infection

We defined two strategies to identify SARS-CoV-2 infection from the EHR spanning all healthcare settings, the first based on diagnostic codes and the second based on laboratory testing. Our first approach relied on the identification of the specific COVID-19 ICD-10-CM diagnosis code U07.1 within the clinical record. We used the ICD-10-CM code as opposed to the corresponding SNOMED codes in the standard OMOP vocabulary given the wider use of ICD-10-CM in administrative data. The U07.1 code, which was introduced on April 1, 2020, was used to define SARS-CoV-2 when used either as (1) a principal diagnosis, or a (2) a principal or a secondary diagnosis of COVID-19 during any healthcare encounter. The principal diagnosis was defined based on the standard OMOP condition status concept code, 32902.[20]

The two diagnosis-based phenotyping strategies were compared to the second approach which was based on the presence of a positive SARS-CoV-2 PCR or antigen test to identify individuals who had documented infection, with manual chart abstraction of samples drawn from discordant subsets to assess the reason for differences. We supplemented this assessment to include potentially related but non-specific diagnoses for severe acute respiratory syndrome (SARS) or coronavirus disease (COVID-19-related diagnoses) based on a subset of codes identified within the National COVID Cohort Collaborative (N3C) phenotype (see **eTable 1** in the Online Supplement).[21]

#### COVID-19 hospitalization

We defined COVID-19 hospitalizations using two strategies. The first identified all hospitalizations with a principal diagnosis of COVID-19 (U07.1). In addition, we defined a second strategy that included individuals with a secondary diagnosis of COVID-19, but with a clinical presentation that was consistent with severe manifestations of COVID-19 defined by a principal diagnosis for acute respiratory failure, pneumonia or sepsis. This approach focused on hospitalizations that were due to COVID-19 rather than incidentally associated with a positive test for the disease during admission for an unrelated diagnosis. The principal diagnoses used in this approach are included in **eTable 2**. There was only a single hospitalization with a diagnosis code J12.82 that has been suggested to identify COVID-19 pneumonia,[21] and was not included in the analysis. Further, to assess validity of diagnosis codes to identify COVID-19 hospitalizations, we compared hospitalizations with COVID-19 diagnosis codes against positive SARS-CoV-2 testing 2 weeks before hospitalization through any time before hospital discharge.

### Study Covariates

We defined key demographic characteristics for individuals, including age, sex, race and ethnicity. Age was defined as completed years on the day of admission, computed from their date of birth. Sex, race and ethnicity were based on what was documented in the medical record. To evaluate the effect of coding strategies on case identification among racial and ethnic minorities, we combined racial/ethnic groups into mutually exclusive groups of Hispanic, non-Hispanic White, non-Hispanic Black, and other race/ethnicity groups.[22 23]

### Study Outcome

Among patients hospitalized with COVID-19, we evaluated differences in in-hospital mortality across case identification strategies. In-hospital mortality was defined based on the discharge disposition of the index (first) COVID-19 hospitalization. Consistent with other studies,[24-26] we used a composite endpoint of in-hospital mortality, transfer to inpatient hospice, or discharge to facility or home-based hospice to define our composite outcome.

### Statistical Analyses

We compared differences in demographic characteristics using the chi-square test for categorical variables and t-test for continuous variables. To assess the performance of COVID-19 diagnoses to accurately identify cases of SARS-CoV-2 infection, we assessed 3 key performance measures: precision (positive predictive value), recall (or sensitivity), and area under the precision recall curve (AUPRC).[27 28]

Analyses were conducted using Spark 2.3.2, Python 3.6.9, and R 3.8. All statistical tests were 2-tailed with a level of significance set at 0.05.

### Manual Chart Abstraction and Validation

Manual chart abstraction was conducted by 2 clinicians independently (RK and WLS) and focused on a sample of randomly selected charts where the diagnosis codes were discordant from laboratory results. For SARS-CoV-2 infections, 10 patient charts were randomly selected from each of the following categories (total 30): (1) principal diagnosis of COVID-19, but negative laboratory diagnosis, (2) secondary diagnosis of COVID-19 but negative laboratory diagnosis, and (3) a positive laboratory diagnosis of SARS-CoV-2 without a corresponding diagnosis code. Furthermore, 10 additional charts were selected for patients who were hospitalized with a principal diagnosis COVID-19 and negative laboratory results, and the clinical documentation was qualitatively reviewed to evaluate the reason for the discrepancy.

### Generalizability of Phenotypes at Mayo Clinic

We constructed equivalent patient cohorts across the 3 Mayo Clinic sites in Minnesota, Arizona, and Florida using the same cohort definitions as outlined in the primary analyses. In these cohorts, we evaluated both the accuracy of the coding strategies in identifying infections with SARS-CoV-2 across care settings, and COVID-19 hospitalizations.

## RESULTS

### SARS-CoV-2 Testing and Diagnosis Rates

There were 69,423 individuals with either a diagnosis of COVID-19 or a positive PCR for SARS-CoV-2 infection at Yale between April 1, 2020 and March 1, 2021. During this period, there were 75,748 SARS-CoV2 infections identified across the 3 Mayo Clinic sites. At Yale, the mean age of patients was 46.0 (±22.4) years and 45.0% of patients were men. Nearly one fourth of patients were of Hispanic ethnicity (22.8%), 57.6% of patients had a recorded race of White and 15.2% were Black (**Table 1**). In contrast, patients in the Mayo Clinic were younger (mean age, 41.8 ± 20.6 years) and were predominantly White (80.6% vs 4.0% Black) (**Table 1**).

**Table 1:** Characteristics of patients across mutually exclusive computable phenotypes from the Yale New-Haven Health System and Mayo Clinic.

| Characteristics | Overall | Principal diagnosis PLUS PCR/Antigen+ | Secondary PLUS PCR/Antigen+ | Principal diagnosis only | Secondary diagnosis only | PCR/Antigen+ only |
| --- | --- | --- | --- | --- | --- | --- |
| **Yale New Haven Health System** |  |  |  |  |  |  |
| **Number of Patients** | 69423 | 38506 | 3449 | 13034 | 6034 | 8400 |
| **Age (mean (SD))** | 46.0 (22.4) | 44.07 (22.3) | 51.2 (23.8) | 49.6 (21.2) | 52.4 (24.6) | 42.6 (20.7) |
| **Men, n (%)** | 31271 (45.0) | 17671 (45.9) | 1629 (47.2) | 5512 (42.3) | 2823 (46.8) | 3636 (43.3) |
| **Race, n (%)** |  |  |  |  |  |  |
| Black | 10582 (15.2) | 6569 (17.1) | 650 (18.8) | 1460 (11.2) | 732 (12.1) | 1171 (13.9) |
| White | 39976 (57.6) | 20665 (53.7) | 1797 (52.1) | 8973 (68.8) | 4221 (70.0) | 4320 (51.4) |
| Asian | 1248 (1.8) | 679 (1.8) | 53 (1.5) | 285 (2.2) | 87 (1.4) | 144 (1.7) |
| Native Hawaiian/Other Pacific Islander | 242 (0.3) | 138 (0.4) | 13 (0.4) | 43 (0.3) | 11 (0.2) | 37 (0.4) |
| American Indian or Alaska Native | 144 (0.2) | 73 (0.2) | 10 (0.3) | 31 (0.2) | 10 (0.2) | 20 (0.2) |
| Other Race | 11833 (17.0) | 7418 (19.3) | 789 (22.9) | 1421 (10.9) | 619 (10.3) | 1586 (18.9) |
| Unknown | 5398 (7.8) | 2964 (7.7) | 137 (4.0) | 821 (6.3) | 354 (5.9) | 1122 (13.4) |
| **Hispanic Ethnicity (%)** | 15829 (22.8) | 10071 (26.2) | 966 (28.0) | 1882 (14.4) | 838 (13.9) | 2072 (24.7) |
| **Mayo Clinic (all 3 sites)** |  |  |  |  |  |  |
| **Number of Patients** | 75748 | 45151 | 1371 | 2069 | 386 | 26771 |
| **Age (mean (SD))** | 41.8 (20.6) | 45.5 (21.0) | 54.8 (22.2) | 57.5 (20.7) | 58.2 (22.3) | 33.5 (16.4) |
| **Men, n (%)** | 37340 (49.3) | 21736 (48.1) | 739 (53.9) | 1025 (49.5) | 200 (51.8) | 13640 (51.0) |
| **Race, n (%)** |  |  |  |  |  |  |
| Black | 3064 (4.0) | 2303 (5.1) | 92 (6.7) | 81 (3.9) | 29 (4.9) | 569 (2.1) |
| White | 61063 (80.6) | 36032 (79.8) | 1129 (82.4) | 1743 (84.2) | 327 (84.7) | 21832 (81.6) |
| Asian | 1685 (2.2) | 1183 (2.6) | 35 (2.6) | 50 (2.4) | <10 (1.6) | 411 (1.5) |
| Native Hawaiian/Other Pacific Islander | 115 (0.2) | 85 (0.2) | <10 (0.1) | <10 (0.2) | <10 (0.8) | 22 (0.1) |
| American Indian or Alaska Native | 353 (0.5) | 217 (0.5) | 32 (2.3) | 45 (2.2) | <10 (1.6) | 53 (0.2) |
| Other Race | 3177 (4.2) | 2353 (5.2) | 50 (3.7) | 63 (3.0) | 15 (3.9) | 696 (2.6) |
| Unknown | 6291 (8.3) | 2978 (6.6) | 32 (2.3) | 83 (4.0) | 10 (2.6) | 3188 (11.9) |
| **Hispanic Ethnicity (%)** | 6057 (8.0) | 4481 (9.9) | 122 (8.9) | 182 (8.8) | 34 (8.8) | 1238 (4.6) |

### Computable Phenotype Accuracy for SARS-CoV-2 Infection at Yale

Of the 69,423 individuals included in our Yale cohort, 51,540 (74.2%) had a principal diagnosis of COVID-19 in the EHR, 61,023 (87.9%) had a principal or a secondary diagnosis, and 50,355 (72.7%) had a positive SARS-CoV-2 PCR or antigen test. There were consistent differences in number of SARS-CoV-2 infections based on the diagnosis and laboratory-based phenotyping strategies throughout the study period, with diagnostic codes being more common than positive laboratory test findings (**Figure 1**). Similar patterns were observed for non-specific coronavirus diagnoses (**eTable 3** in the Online Supplement).

**Figure 1:**
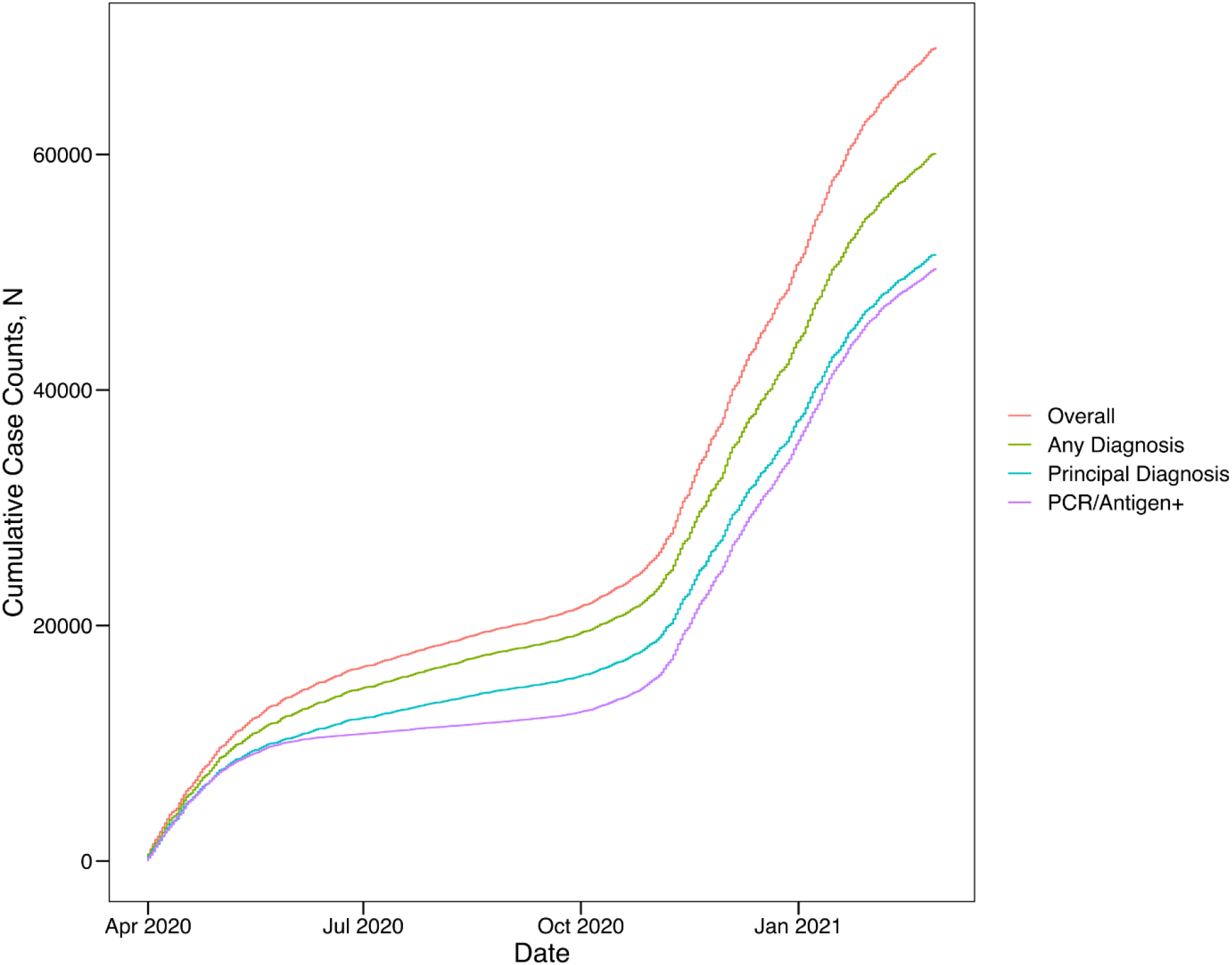
Cumulative SARS-CoV-2 cases by adjudication strategy across the study period.

Of the 50,355 patients with a positive laboratory test for SARS-CoV-2, only 38,506 (76.5%) had a principal diagnosis and an additional 3449 (6.8%) had a secondary diagnosis of COVID-19 recorded in the EHR. The remaining 8400 (16.7%) had no COVID-19 diagnosis recorded as a principal or secondary diagnostic code within the medical record (**Figure 2**). Moreover, there were 19,068 patients (31.2%) who had a principal or a secondary COVID-19 diagnosis without a positive lab test for SARS-CoV-2. The characteristics of patients in these groups are included in **Table 1**.

**Figure 2:**
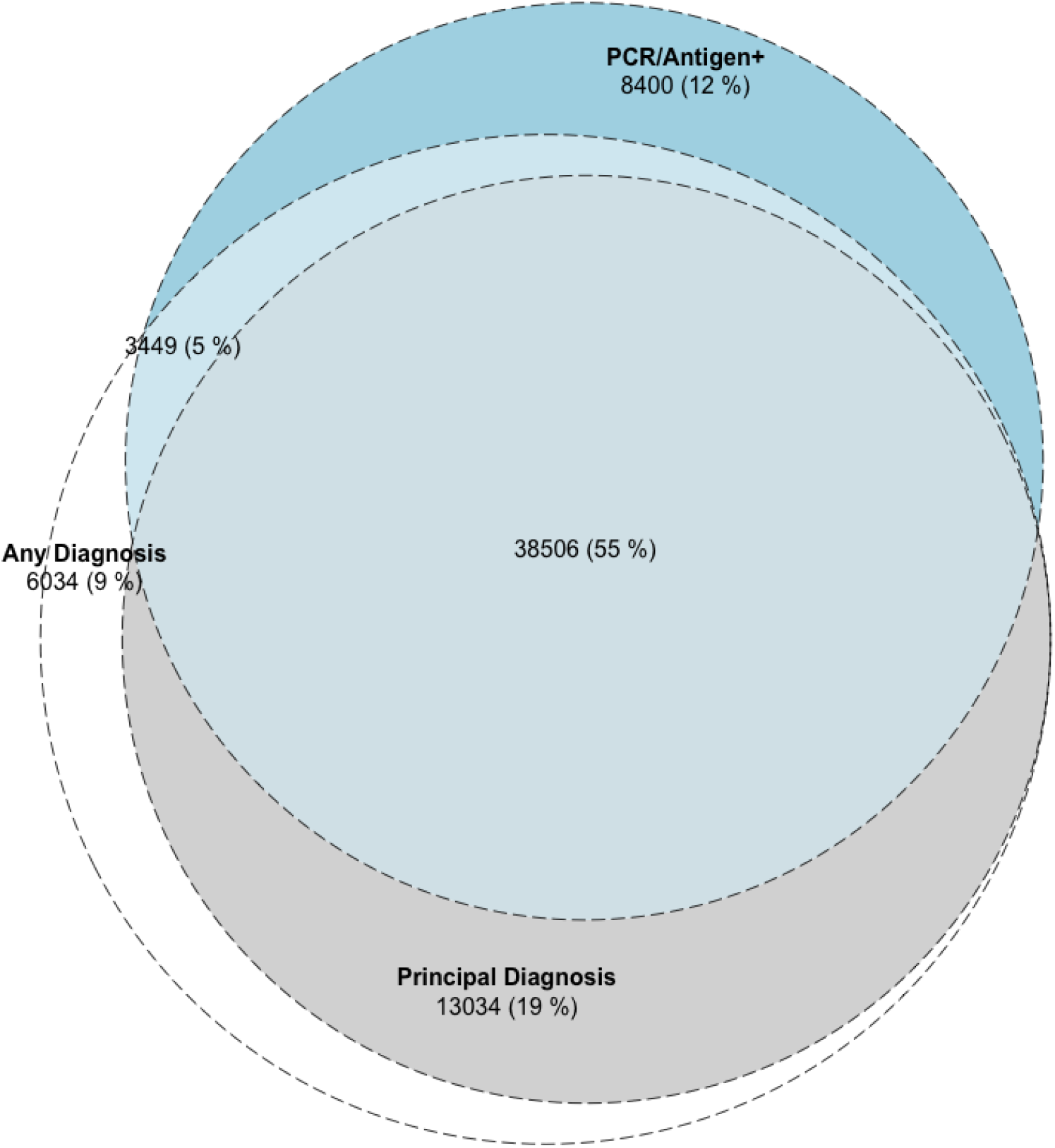
Computable phenotypes for SARS-CoV-2 infection across the study period at Yale New Haven Health System

In a manual chart review of a random sample of 30 patients with a diagnosis and without a positive SARS-CoV-2 test, all had a healthcare visit for SARS-CoV-2 testing but with a subsequent negative laboratory test.

The use of a principal diagnosis code of COVID-19 as the criteria to identify SARS-CoV-2 infection had a precision (or positive predictive value) of 74.7% (95% CI, 74.3% to 75.1%) and a recall (or sensitivity) of 76.5% (95% CI, 76.1% to 76.8%) with an AUPRC of 0.17. Inferring infection with any diagnosis code for COVID-19 had worse performance characteristics (precision: 68.8% [68.4% to 69.1%]), recall: 83.3% [83.0% to 83.6%]), and AUPRC 0.12).

There were significant differences in concordance between patient identification strategies during the study period (**eFigure 1**). Among patients with either a diagnosis code for COVID-19 or a laboratory diagnosis of SARS-CoV-2, 51% of patients had both a diagnosis code and a positive laboratory test between April and August 2020, while 65% of patients had both present between September 2020 and March 2021 (P<0.001). There were significant differences across racial and ethnic groups (**Figure 3A**). Among Hispanics and non-Hispanic Black patients, 69.5% and 68.7% patients, respectively had both a diagnosis of COVID-19 and a positive laboratory test, compared with 54.5% of non-Hispanic White patients (P<0.001). There was also a significant difference by sex, with more women having a concomitant diagnosis code and positive laboratory test than men (61.4% vs 59.1%, P<0.001) (**Figure 3B**).

**Figure 3:**
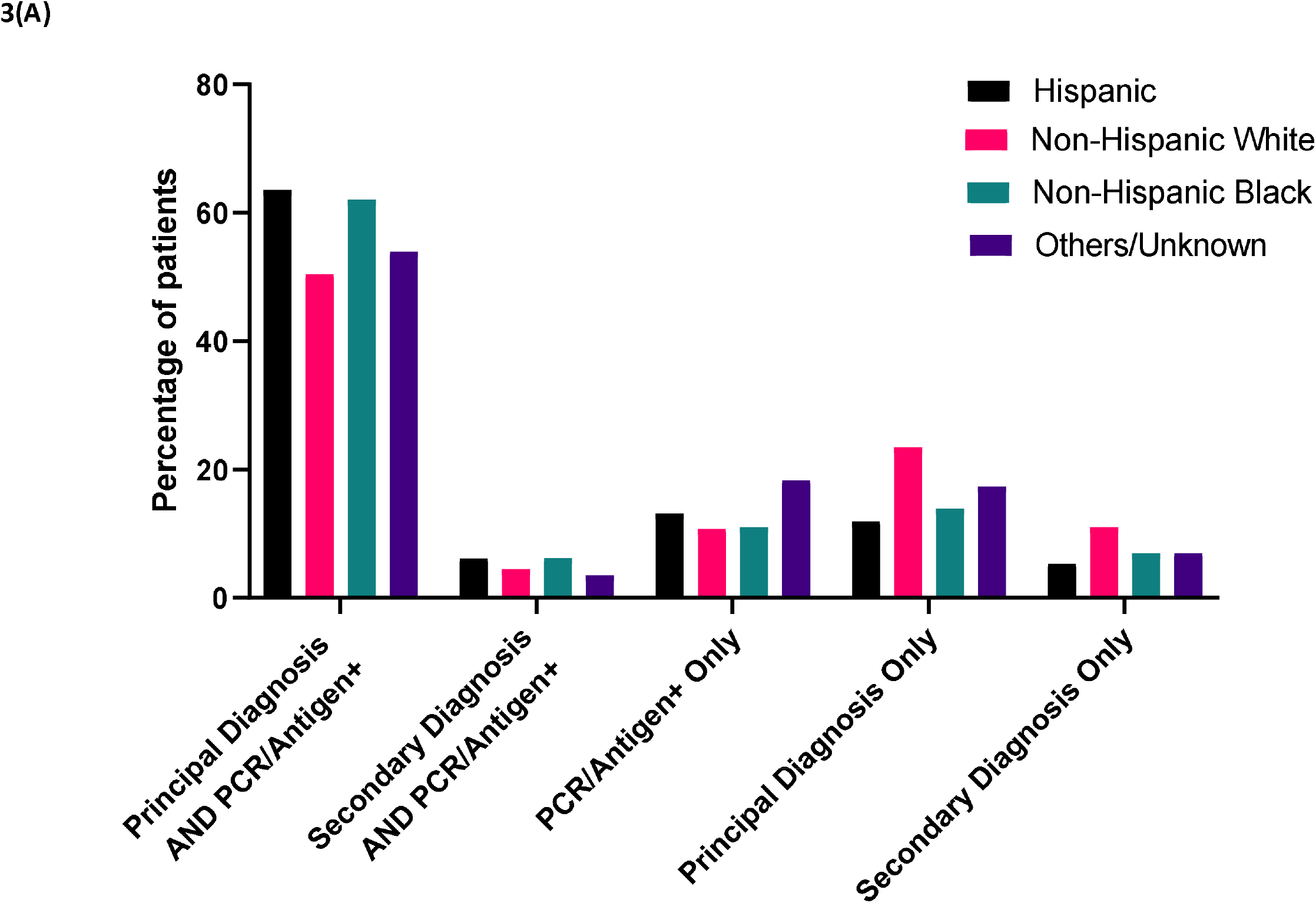
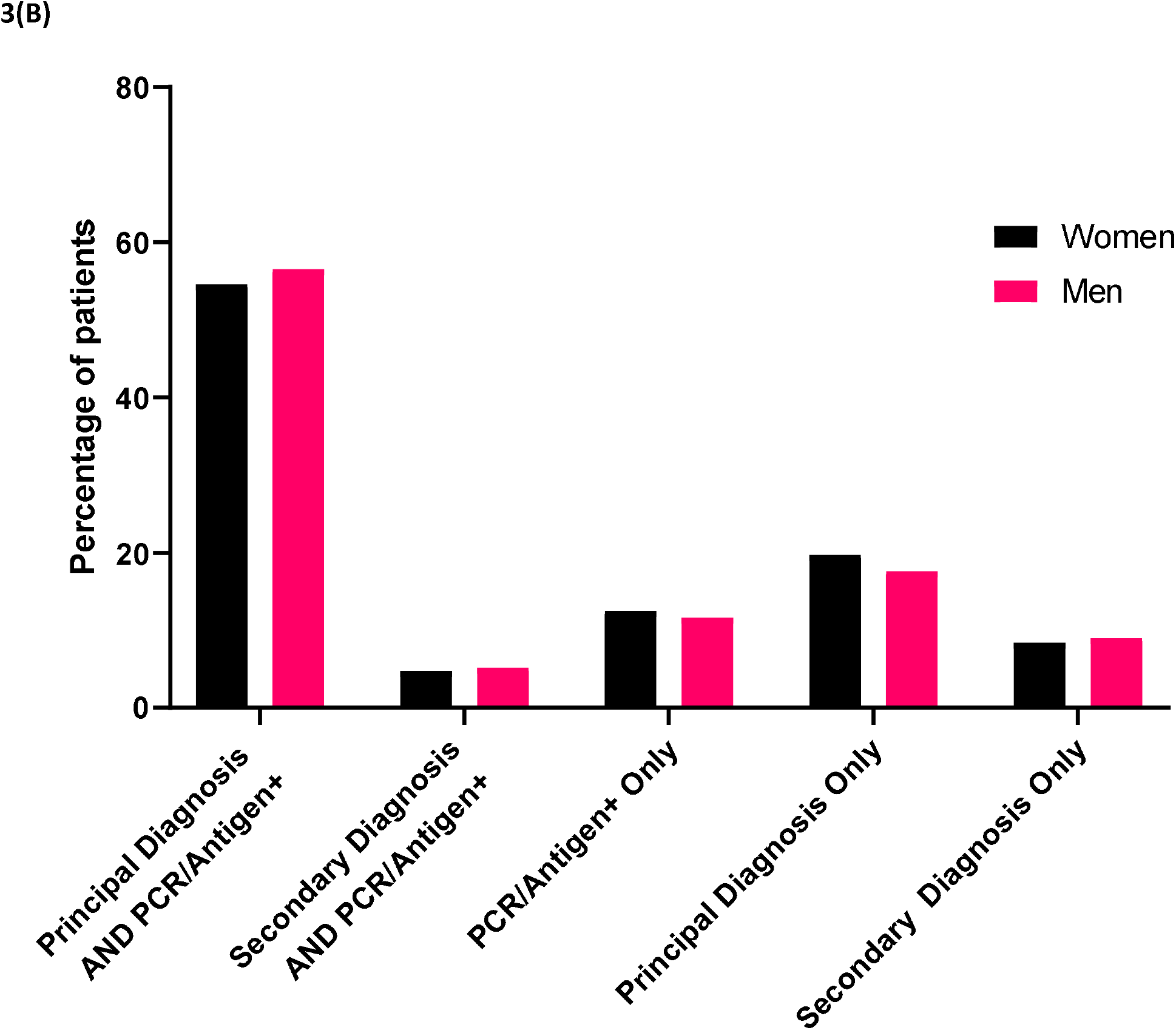
Computable phenotypes for SARS-CoV-2 infection by (A) Race/Ethnicity and (B) Sex in the Yale-New Haven Health System.

### Accuracy of Phenotypes for SARS-CoV-2 Infections Across Mayo Clinic Sites

At Mayo Clinic, both a principal diagnosis of COVID-19 or a principal or secondary of COVID-19 were associated with high precision for SARS-CoV2 infection (95.6% for principal diagnosis, and 95.0% for secondary diagnosis, **Figure 4**). However, the recall (or sensitivity) was low with both the principal (63.0%) or any diagnosis of COVID-19 (63.5%). Further, there was substantial variation across the Mayo Clinic sites, with the sensitivity of a COVID-19 diagnosis identifying SARS-CoV-2 infection varying between 59.2% in Rochester to 97.3% in Arizona (**Figure 4**).

**Figure 4:**
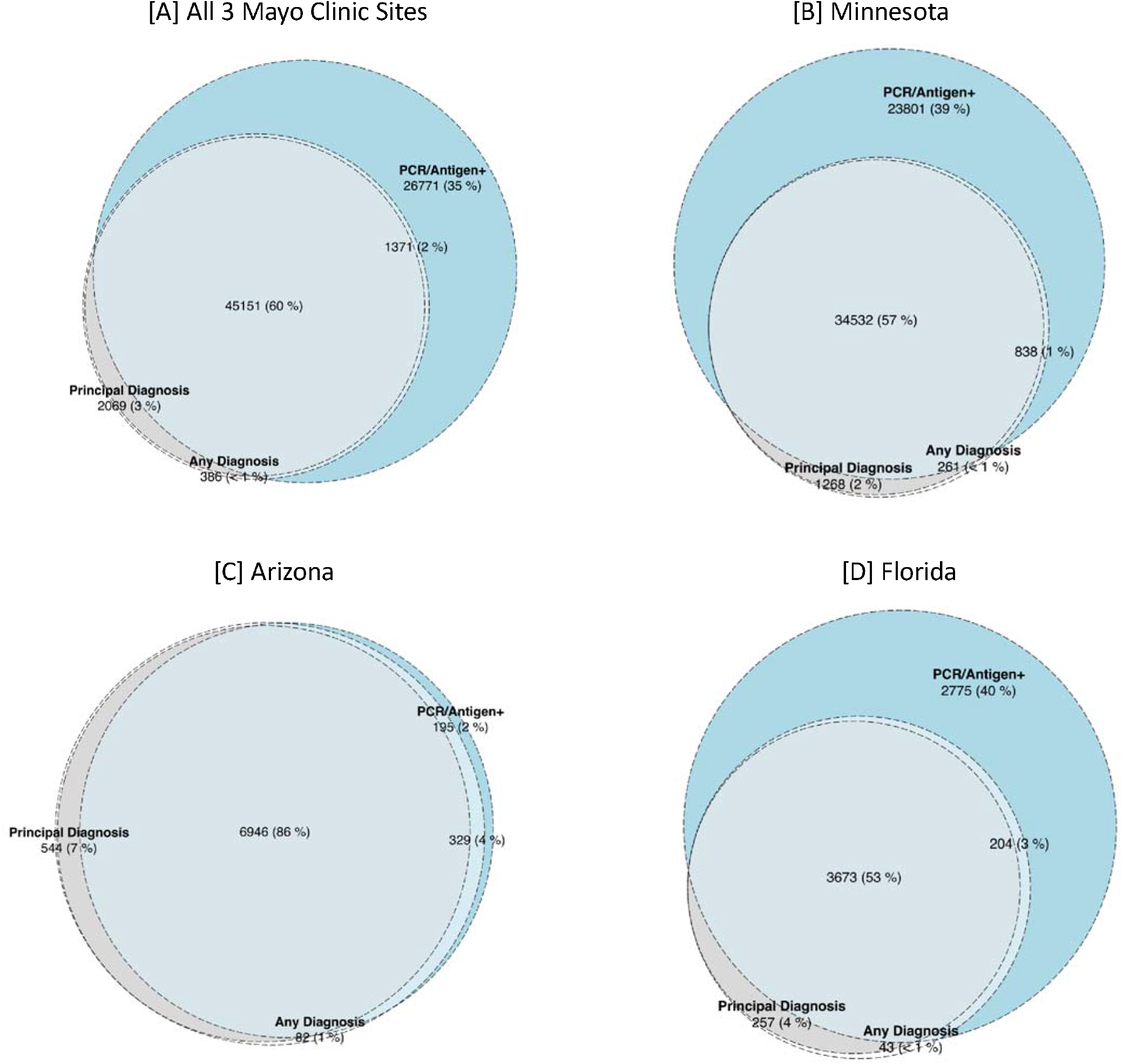
Computable phenotypes for SARS-CoV-2 infection across the study period at the Mayo Clinic System

### Computable Phenotype Accuracy for COVID-19 Hospitalization

Based on visit start date, there were a total of 5,555 discharges using our overall phenotyping strategy from April 1, 2020 and January 31, 2021 at Yale. Of these, 5,109 (92.0%) discharges had a principal diagnosis of COVID-19 and the remaining 446 had a principal diagnosis for a COVID-19 related severe presentation and a secondary diagnosis of COVID-19 on the same visit. Finally, there were 343 individuals who had a secondary, but not primary, diagnosis of COVID-19 which were excluded from analysis as these diagnoses were incidental findings or hospital-acquired infections. Those with a COVID-19 primary diagnosis were less frequently male (50.9% vs 59.6%, P <0.001) and were more frequently Black (20.9% vs 17.3%, P = 0.02).

The vast majority of patients had a positive SARS-CoV-2 PCR or antigen test during their hospitalization across both patients hospitalized with a COVID-19 primary diagnosis (94.8%, n=4843) or a secondary diagnosis (91.9%, n=410). A manually abstracted sample of 10 charts of hospitalized individuals without a positive laboratory test but with a principal diagnosis of COVID-19 found that 7 of these patients had a positive COVID-19 test at another healthcare facility prior to presentation and 3 had a strong clinical suspicion for COVID-19 but a negative PCR test.

### COVID-19 Hospitalization Phenotypes Across Mayo Clinic Sites

A smaller proportion of patients with a principal diagnosis of COVID-19 in the Mayo Clinic System, as compared with those at Yale, had a positive SARS-CoV-2 PCR or antigen test during the hospitalization (80.5%, n=2,378), but the proportion was similar between Mayo Clinic and Yale for patients with a secondary diagnosis (90.7%, 331). A manually abstracted sample of 10 charts among individuals who had a principal diagnosis of COVID-19 without a positive laboratory test identified that 9 of these patients had a positive SARS-CoV-2 test at another healthcare facility prior to presentation and 1 did not have a documented SARS-CoV-2 test.

### Relationship between COVID-19 Hospitalization Phenotype Definition and In-Hospital Mortality Rate

At Yale, the in-hospital mortality rate for those hospitalized with a principal diagnosis of COVID-19 was 13.2% (675 of 5,109), which was significantly lower than those with a secondary diagnosis of COVID-19 and related primary diagnosis code for sepsis or respiratory failure, who had an in-hospital mortality that was nearly double (28.0%, 125 of 446, P<0.001) (**Table 2**). This pattern was also observed at Mayo Clinic, with an 8.0% (237 of 2954) in-hospital mortality rate among patients with a principal diagnosis of COVID-19, compared with 22.7% (83 of 365) among those with a secondary diagnosis of COVID-19. This was observed across all 3 Mayo Clinic sites (**Figure 5**).

**Table 2:** Characteristics of hospitalized COVID-19 patients with a principal or secondary diagnosis of COVID-19 (U07.1).

| Characteristics | Overall | Principal diagnosis of COVID-19 | Secondary diagnosis of COVID-19* |
| --- | --- | --- | --- |
|  | **Yale New Haven Health System** |  |  |
| **Number of Patients** | 5555 | 5109 | 446 |
| **Age (mean (SD))** | 66.37 (17.59) | 66.17 (17.68) | 68.63 (16.44) |
| **Men, n (%)** | 2867 (51.6) | 2601 (50.9) | 266 (59.6) |
| **Race, n (%)** |  |  |  |
| Black | 1145 (20.6) | 1068 (20.9) | 77 (17.3) |
| White | 3156 (56.8) | 2880 (56.4) | 276 (61.9) |
| Asian | 103 (1.9) | 96 (1.9) | <10 (1.6) |
| Native Hawaiian/Other Pacific Islander | 19 (0.3) | 19 (0.4) | 0 (0.0) |
| American Indian or Alaska Native | 12 (0.2) | 11 (0.2) | <10 (0.2) |
| Other Race | 1043 (18.8) | 960 (18.8) | 83 (18.6) |
| Unknown | 77 (1.4) | 75 (1.5) | <10 (0.4) |
| **Hispanic Ethnicity (%)** | 1243 (22.4) | 1152 (22.5) | 91 (20.4) |
| **In-hospital mortality/Discharge to Hospice, n (%)** | 800 (14.4) | 675 (13.2) | 125 (28.0) |
|  | **Mayo Clinic** |  |  |
| **Number of Patients** | 3319 | 2954 | 365 |
| **Age (mean (SD))** | 65.47 (17.84) | 65.41 (17.98) | 65.92 (16.64) |
| **Men, n (%)** | 1893 (57.0) | 1659 (56.6) | 234 (64.1) |
| **Race, n (%)** |  |  |  |
| Black | 173 (5.2) | 149 (5.0) | 24 (6.6) |
| White | 2714 (81.8) | 2427 (82.2) | 287 (76.6) |
| Asian | 110 (3.3) | 93 (3.2) | 17 (4.7) |
| Native Hawaiian/Other Pacific Islander | <10 (0.2) | <10 (0.2) | 0 (0.0) |
| American Indian or Alaska Native | 116 (3.5) | 100 (3.4) | 16 (4.4) |
| Other Race | 126 (3.8) | 109 (3.7) | 17 (4.7) |
| Unknown | 74 (2.2) | 70 (2.4) | 4 (1.1) |
| **Hispanic Ethnicity, n (%)** | 326 (9.8) | 277 (9.4) | 49 (13.4) |
| **In-hospital mortality/Discharge to Hospice, n (%)** | 320 (9.6) | 237 (8.0) | 83 (22.7) |

\*with a principal diagnosis for respiratory failure, sepsis or pneumonia

**Figure 5:**
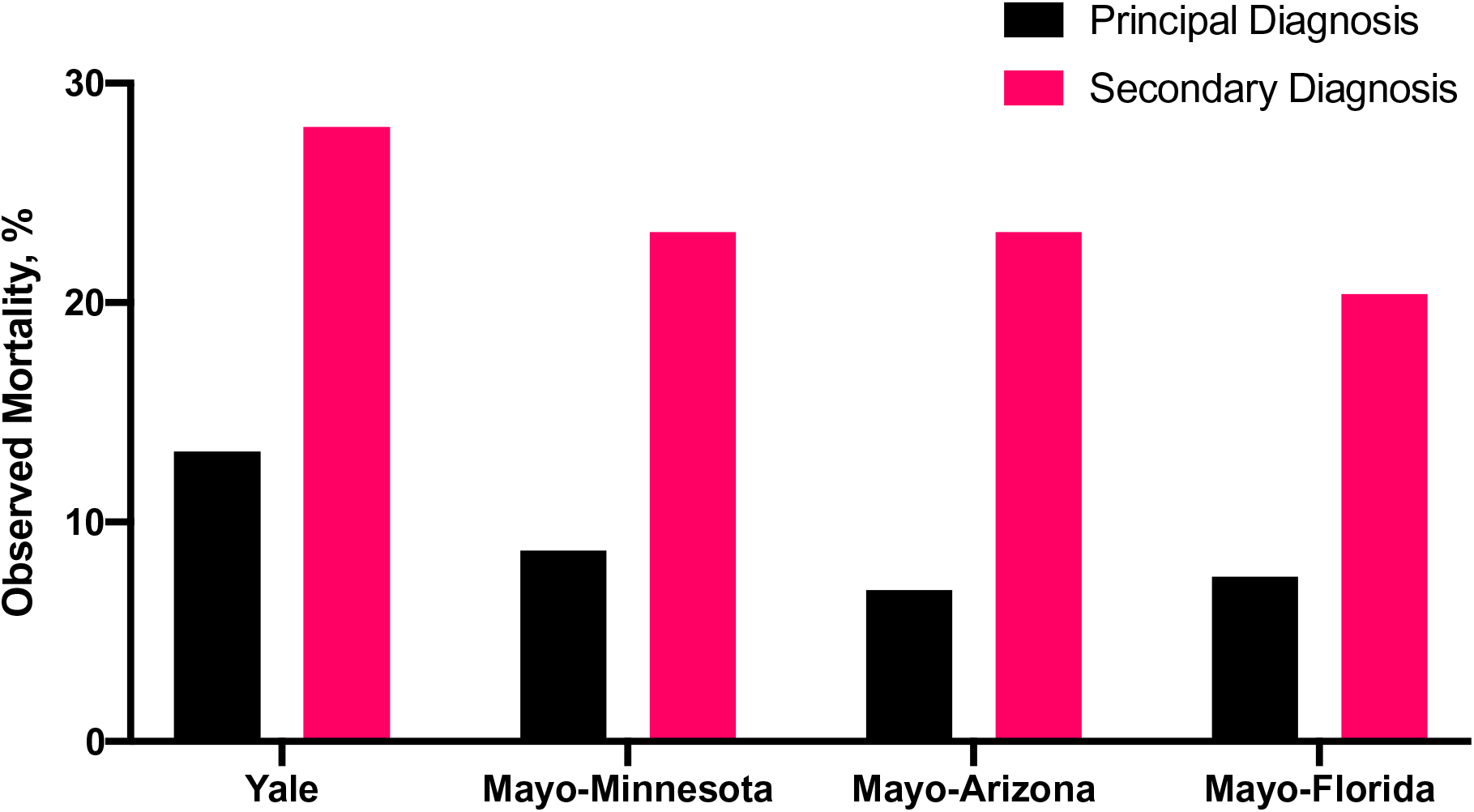
Mortality for COVID-19 hospitalizations defined by principal and secondary diagnosis by study site. Mortality represents in-hospital death and discharge to hospice from index admission.

## DISCUSSION

In two large, integrated health systems with multiple care delivery networks and associated outpatient clinics, COVID-19 diagnosis codes alone were frequently inaccurate for case identification and epidemiological surveillance of SARS-CoV-2 infection, with significant variation across two major health systems. In contrast, a principal diagnosis of COVID-19 diagnosis correctly consistently identified nearly 95% of patients admitted with COVID-19, though would miss an additional 10% of people with a clinical profile consistent with severe COVID-19, but with a secondary diagnosis of COVID-19. Moreover, computable phenotype definitions for hospitalizations had implications for outcome ascertainment, where using only a principal discharge diagnosis of COVID-19 to identify hospitalizations would miss many people in a high-risk group who had an over 2-fold higher mortality rate compared with patients with a principal diagnosis of COVID-19.

This study expands the literature in several key ways. To our knowledge, in the largest study leveraging the EHR as a source of real-world data, rather than administrative claims, to evaluate the accuracy of strategies for SARS-CoV-2 case identification across outpatient and inpatient healthcare settings. Moreover, in addition to identifying the accuracy of computable phenotype definitions, we evaluated the association of phenotype definitions on inferred short-term outcomes. A previous study found that the COVID-19 diagnosis code, U07.1 was rapidly incorporated into the workflow of US hospitals in early 2020,[29] and among hospitalized patients, had a high sensitivity and specificity of the code for laboratory confirmed disease. However, that study was limited in using data only through May 2020, with only 4965 SARS-CoV-2 positive laboratory tests. Moreover, the accuracy measures were driven by the 89.6% of the cohort that did not have either a positive test or a diagnosis code for COVID-19.[29] The evaluation of COVID-19 diagnosis codes also focused exclusively among hospitalized patients and did not evaluate the role of diagnosis codes in case surveillance. We confirm that an inpatient diagnosis of COVID-19 has retained a large positive predictive value for clinical COVID-19. Yet, we found significant heterogeneity in outcomes based on whether COVID-19 was included as a principal or a secondary diagnosis. Finally, we evaluated the approach across 2 different hospital systems spanning 4 distinct geographic regions over an entire year.

There are many possible reasons for the incorrect classification of SARS-CoV-2 infections by diagnosis code. Many studies have shown the apparent inaccuracy of various EHR data elements, such as the clinical history and problem list.[30 31] Clinical uncertainty related to a diagnosis, potential stigma associated with the addition of a diagnosis to the medical record, clinical workflows that do not promote the capture of structured data elements, and miscoded diagnoses can all impact the ability to define a digital phenotype that accurately identifies patients.[30 32 33] Moreover, diagnosis codes are often included when evaluating a suspected condition and may be misconstrued as proof of diagnosis, particularly in data captured in near real-time.

We also gained additional insights into the mechanism behind the discordance between diagnosis codes and laboratory results through manual chart review. This allowed us to carefully define a cohort of patients with COVID-19 based on clinical definition. Moreover, it also allowed us to exclude patients who did not have an admission for COVID-19 despite an incidental positive SARS-CoV-2 test and secondary diagnosis, as during the pandemic many clinical presentations (such as presentations for trauma) did have an positive SARS-CoV-2 screening for placement and isolation without clinical disease.[3] Our racially and ethnically diverse study population also allowed our study to uncover patterns of differential performance of diagnosis codes for case surveillance in racial/ethnic minority groups, in whom the presence of diagnosis codes more accurately aligned with laboratory confirmed disease.

Our study highlights the value of health information systems in disease monitoring through logical cohort definitions, which can be explicitly confirmed across different data elements. Moreover, our work supports the need for continuous monitoring and validation of computable phenotypes, especially those that rely solely on diagnosis codes, such as those used to analyze administrative data sets.

Another unique aspect of our approach is the availability of all EHR data in the OMOP CDM at Yale that is updated daily, allowing rapid and serially updated assessment of such cohort definitions and disease surveillance over the course of the pandemic, allowing the most up-to-date assessment of specific patient populations in a rapidly evolving condition and their outcomes. A similar common data model available across health systems could allow rapid investigation of generalizable disease phenotypes, offering the ability to assess valid case definitions for large scale real-world studies.

Our study has several limitations. First, while we focused on two broad interconnected health systems and affiliated laboratories and receive testing information from laboratories that exchange data via the Epic EHR, not all external laboratory data were not available from testing in the outpatient setting. However, in manual chart review of a sample of patients with an outpatient diagnosis of COVID-19 without a reported positive PCR or antigen test, all such records were for patients undergoing SARS-CoV-2 testing with the diagnosis assigned for the clinical or laboratory encounter to obtain the test. Second, we cannot infer coding practices at other institutions not included in the study. However, the two large integrated multi-hospital health systems included in the study demonstrated substantial inter-hospital heterogeneity in coding practices. Such a site-to-site variation is likely prevalent across hospitals not included in the study, making the use of diagnosis codes not a reliable measure for SARS-CoV-2 infection surveillance.

## CONCLUSIONS

The use of COVID-19 diagnosis codes misclassified SARS-CoV-2 infection status for many people, with implications for clinical research and epidemiological surveillance. Moreover, the codes had different performance across two academic health systems and identified groups with different risks of mortality. Data from the EHR can be used to in conjunction with diagnosis codes to improve the identification of people hospitalized with COVID-19.

## Data Availability

Patient-level data represented protected health information and cannot be shared. Summary data for the information presented in the figures are available upon request.

