## Supplementary material for "Accuracy of Computable Phenotyping Approaches for SARS-CoV-2 Infection and COVID-19 Hospitalizations from the Electronic Health Record": Online Supplement

**eTable 1: Non-specific coronavirus diagnosis codes.**

| **ICD-10-CM Code** | **Description** |
| --- | --- |
| B97.21 | SARS-associated coronavirus as the cause of diseases classified elsewhere |
| B97.29 | Other coronavirus as the cause of diseases classified elsewhere |
| B34.2 | Coronavirus infection, unspecified |
| J12.81 | Pneumonia due to SARS-associated coronavirus |

*Codes included in the initial cohort selection strategy by the National COVID Cohort Collaborative (N3C). Abbreviations: ICD-10-CM: International Classification of Diseases, Tenth Revision, Clinical Modification; SARS – severe acute respiratory syndrome.

**eTable 2:** Primary diagnosis codes for severe COVID-19 manifestations or non-specific coronavirus infection recorded in our study population with a secondary diagnosis of COVID-19 (U07.1).

| **ICD-10-CM Code** | **Description** |
| --- | --- |
| A41.89 | Other specified sepsis |
| A41.9 | Sepsis, unspecified organism |
| B34.2 | Coronavirus infection, unspecified |
| J12.89 | Other viral pneumonia |
| J18.9 | Pneumonia, unspecified organism |
| J80 | Acute respiratory distress syndrome |
| J96.01 | Acute respiratory failure with hypoxia |
| J96.21 | Acute and chronic respiratory failure with hypoxia |

Abbreviation: ICD-10-CM: International Classification of Diseases, Tenth Revision, Clinical Modification

**eTable 3:** Monthly counts of non-specific coronavirus associated diagnoses and those with a laboratory diagnosis in the Yale New Haven Health System (i.e. positive laboratory test for SARS-CoV-2 nucleic acid or antigen).

|  | **2020** | | | | | | | | | **2021** | |
| --- | --- | --- | --- | --- | --- | --- | --- | --- | --- | --- | --- |
|  | **Apr** | **May** | **Jun** | **Jul** | **Aug** | **Sep** | **Oct** | **Nov** | **Dec** | **Jan** | **Feb** |
| B97.21 | 29 | 16 | 3 | 17 | 4 | 4 | 14 | 15 | 16 | 12 | 3 |
| B97.29 | 146 | 172 | 159 | 206 | 139 | 87 | 49 | 90 | 64 | 25 | 6 |
| B34.2 | 205 | 67 | 40 | 86 | 64 | 68 | 96 | 338 | 200 | 52 | 15 |
| J12.81 | 37 | 33 | 11 | 3 | 2 | 3 | 5 | 12 | 20 | 25 | 11 |
| B97.27 + Lab diagnosis | 9 | 9 | 1 | 3 | 1 | 1 | 4 | 2 | 5 | 4 | 1 |
| B97.29 + Lab diagnosis | 43 | 29 | 10 | 12 | 21 | 16 | 11 | 51 | 22 | 8 | 2 |
| B34.2 + Lab diagnosis | 75 | 21 | 9 | 5 | 6 | 3 | 4 | 61 | 63 | 19 | 3 |
| J12.81 + Lab diagnosis | 20 | 15 | 6 | 1 | 1 | 3 | 2 | 7 | 11 | 16 | 8 |

**eFigure 1:** Diagnostic groups for SARS-CoV2 infection across the study period, (A) April 2020 to August 2020, (B) September 2020 to March 2021

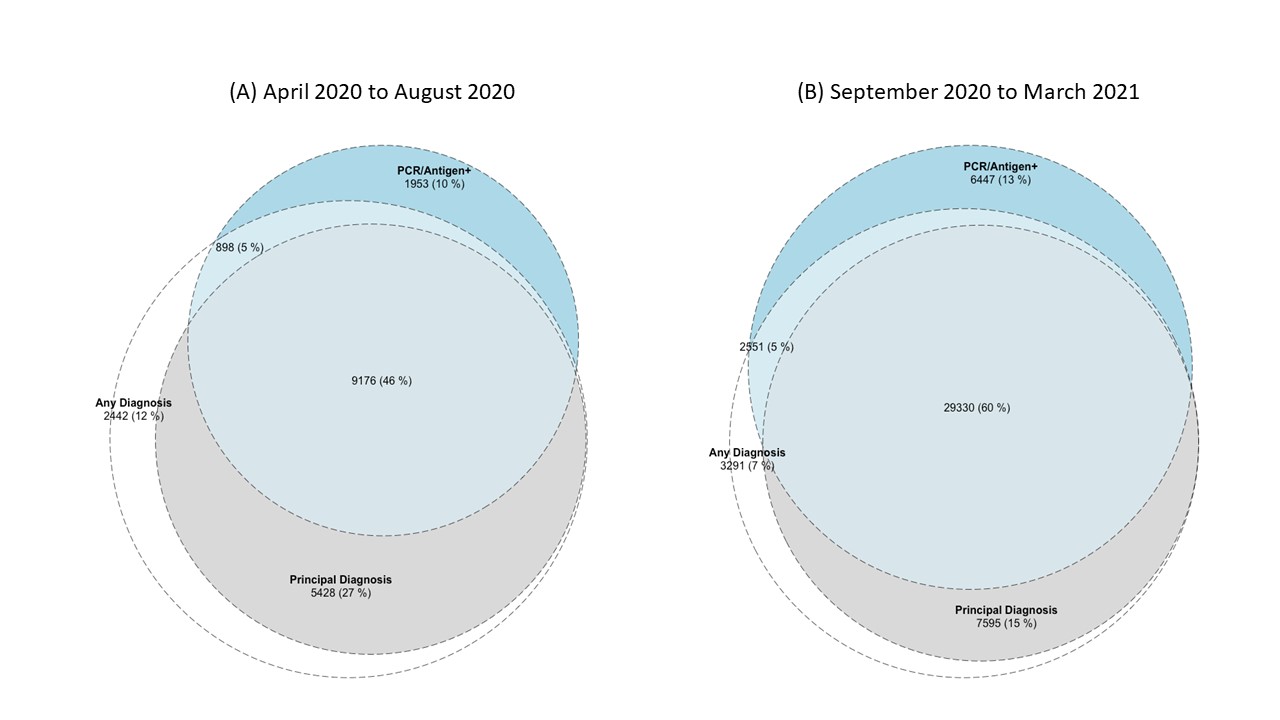
